# Patient and provider perspectives on factors that influence the implementation of a community and hospital care bundle to improve the treatment of patients with peripheral arterial disease in primary and secondary care

**DOI:** 10.1101/2022.07.18.22277752

**Authors:** Clair Le Boutillier, Athanasios Saratzis, Prakash Saha, Ruth Benson, Bernadeta Bridgwood, Emma Watson, Vanessa Lawrence

## Abstract

**Background:** The Community and Hospital cAre Bundle to improve the medical treatment of cLaudIcation and critical limb iSchaemia (CHABLIS) study is a prospective mixed-methods study across NHS hospitals and primary care networks, which aims to determine the feasibility of using a complex intervention in the form of a care bundle, consisting of checklists, leaflets and letters, called the LEGS intervention (LEaflet Gp letter Structured checklist), to improve the care of patients with peripheral arterial disease (PAD). The aim of this qualitative study was to gain an understanding of the acceptability of the provision and delivery of the LEGS intervention, by patients, general practitioners and secondary care clinicians. Engaging stakeholders in these conversations provides insights for future intervention refinement, uptake and implementation.

**Methods:** This qualitative study was embedded within the CHABLIS study. Twenty-five semi-structured telephone interviews were conducted with i) patients who had received the intervention (n=11), ii) secondary care clinicians responsible for delivering the intervention (n=8), and iii) general practitioners (n=6). Data were initially analysed using inductive descriptive thematic analysis. The consolidated framework for implementation research was then used as a matrix to explore patterns in the data and to map connections between the three participant groups. Lastly, interpretive analysis allowed for refining, and a final coding frame was developed.

**Results:** Four overarching themes were identified: i) The potential to make a difference, ii) A solution to address the gap in no man’s land, iii), Prioritising and making it happen and iv) Personalised information and supportive conversations for taking on the advice. The intervention was viewed as an opportunity to meet patient needs, and to develop shared primary and secondary care working practices. The impetus for prioritising and delivering the intervention was further driven by its flexibility and adaptability to be tailored to the individual and to the environment.

**Conclusions:** The LEGS intervention can be tailored for use at early and late stages of PAD, can be provided across primary and secondary care settings, and provides an opportunity to promote shared working across the primary-secondary care interface.

**Contributions to the literature:**

- Primary and secondary care providers acknowledged the need for an intervention to support them to deliver guideline-based PAD treatment, and to target the intervention earlier in the PAD treatment pathway.
- A gap was identified in terms of support for patients and providers between the time of diagnosis of early-stage PAD (e.g., claudication) and a subsequent potential diagnosis of advanced PAD. The LEGS intervention can be used to fill this gap by enabling providers to support patients to receive help, education, support, or appropriate medication to address their condition.
- Patient-provider interactions that promote shared decision-making and that support patient preference are also important determinants in the success of implementation.

## Background

Peripheral Arterial Disease (PAD) affects a fifth of people over the age of sixty in the United Kingdom. This prevalence is also expected to rise due to sedentary lifestyles, poor dietary choices, a longer life-expectancy, and rising numbers of those with diabetes (Cea-Soriano et al., 2018; Herrington et al., 2016). Patients with PAD who develop symptoms present either with Intermittent Claudication (IC) or in some cases, Chronic Limb Threatening Ischaemia (CLTI) where the reduction in blood flow is so severe that it causes pain at rest, ulceration, or gangrene (Dhaliwal & Mukherjee, 2007). Patients with IC are more likely to suffer a cardiovascular event compared to age-matched individuals, and the one-year risk of limb amputation for those with CLTI is 30%. More than half of those diagnosed with symptomatic PAD are expected to die, have an amputation or a major cardiovascular event within five years (Dua & Lee, 2016; Morley et al., 2018; Nehler et al., 2014; Parvar et al., 2018).

The treatment of these high-risk individuals is shared between primary and secondary care. While established national and international guidance (National Institute for Health and Care Excellence, 2020) outlines how PAD risk-factors should be addressed to prevent cardiac events and amputations, a study conducted by our group found that only a tenth of those diagnosed with PAD were prescribed appropriate preventive medication, and almost none received structured lifestyle or dietary support (Saratzis et al., 2019). As a result, and following patient and public engagement (that is, interviews and workshops with patients and a national consultation with primary and secondary care clinicians) and a review of the existing evidence, a complex intervention in the form of a care bundle was developed to support patients, General Practitioners (GPs), and secondary care clinicians to actively manage cardiovascular risk-factors, and to improve implementation of existing guidance when treating patients with PAD. The care bundle, called the LEGS intervention (Leaflet, Gp letter, Structured checklist) has three elements: i) a one-page clinical checklists for the ward and a separate one-page clinical checklist for the secondary care clinic, ii) a patient and carer education leaflet (developed with patients), iii) a GP action letter, that can be sent to GPs at regular intervals to inform them of the PAD diagnosis and corresponding practice guidance. The LEGS intervention and full study protocol are available online (isrctn.com/ISRCTN13202085).

This qualitative study was embedded within the Community and Hospital cAre Bundle to improve the medical treatment of cLaudIcation and critical limb iSchaemia (CHABLIS) study. CHABLIS is a multicentre prospective mixed-methods cohort study that aims to determine the feasibility and effectiveness of using a PAD care bundle, called the LEGS intervention (LEaflet Gp letter Structured checklist). The aim of the qualitative study was to gain an understanding of the acceptability of the LEGS intervention, by exploring patient and provider perspectives on receiving and delivering the intervention (Skivington et al., 2021). The objectives of the qualitative study were to 1) to explore patient perspectives on receiving the LEGS intervention, 2) to gather secondary care clinician views on acceptability and usability, and to understand factors that influence the success of implementation, and 3) to identify barriers and facilitators that influence the success of future implementation from the GP perspective.

## METHODS

### Study design

The study used qualitative methods to provide insights into the acceptability, usability, and relevance of the intervention as well as the complexities of implementation. Semi-structured individual interviews were conducted with patients and providers to gain an understanding of the barriers and facilitators to implementation in secondary and primary care.

To meet objective 1, semi-structured individual interviews were conducted with patients with symptomatic PAD (i.e., IC, ischaemic rest pain, or CTLI), who had received the intervention (either in an outpatient clinic or as a part of inpatient treatment). To meet objective 2, semi-structured individual interviews were conducted with secondary care clinicians responsible for delivering the intervention, including junior doctors, vascular nurses, and vascular surgeons. Staff and patients were recruited from two implementation sites: University Hospitals of Leicester NHS Trust (UHL) and Guy’s and St Thomas’ NHS Foundation Trust (GSTT). To meet objective 3, semi-structured individual interviews were conducted with GPs to gain an understanding of the barriers and facilitators to future implementation in primary care and across the NHS.

Participants were selected purposively (both those who were receiving or responsible for delivering the LEGS intervention) on the basis that they are the target adopters of behaviours that influence the success of future implementation and/or could offer a particular perspective on implementation. Participants were over 18, and proficient in English. Eligible patients were approached and recruited by the local research team (for example, local principal investigator, research nurse), via face-to-face consultation as a part of outpatient clinic or hospital admission. Potential secondary care clinician participants were approached and recruited by the local research team. GP participants were recruited from Clinical Commissioning Groups linked to the two secondary care implementation sites, chosen to enhance joined up working and to provide a local perspective on implementation across community and hospital settings. Potential GP participants were first approached by the local Clinical Research Network research delivery manager or local contacts, and subsequently recruited by the lead author via telephone or email. Written or verbal informed consent was obtained from each participant after a full explanation and information leaflet was given and time allowed for consideration. The right of each participant to refuse to participate without giving reasons was respected. All participants were free to withdraw from the study at any time without giving reasons and without prejudicing further treatment or employment.

### Data collection

Interviews used flexible open-ended questions for early data collection to gather a rich and detailed understanding of participants’ perspectives. The patient and provider interview schedules were revised iteratively in response to the priorities and concerns of participants and are included in Online Data Supplement (ODS) 1. Interviews were conducted by telephone by the lead author (a qualitative research fellow), lasted around 45 minutes, were audio-recorded and transcribed verbatim. Researcher reflexive notes were kept after each interview to consider the interaction with the participant, and to detail initial thoughts. Interviews were conducted between September and November 2021.

### Data Analysis

Data from the three participant groups (patients, secondary care clinicians and GPs) were first analysed separately using inductive thematic analysis, where analytical concepts and perspectives are generated from the data in a deliberate and systematic way (Braun & Clarke, 2006). Data analysis began with repeated re-reading of individual transcripts and re-listening of sound files for data immersion. This was followed by line-by-line open coding, where extracts were coded under one or several descriptive themes, that is organising the data according to semantic content, to capture their meaning and reflect the content of the data. Each theme was refined, and where data allowed, further sub-themes were developed (CL). The Consolidated Framework for Implementation Research (CFIR) was then used as a matrix to organise the early themes, to explore patterns and relationships in the data, and to map connections between the themes and the three participant groups (Damschroder et al, 2009). Lastly, interpretive analysis allowed for the refining of the specifics of themes and thematic patterns, and a final coding frame for patients and providers was developed. Refinements to the specifics of themes, and thematic patterns continued until a useful and meaningful analysis was achieved (Braun & Clarke, 2021).

Data collection occurred concurrently with data analysis; NVivo QSR International qualitative analysis software (version 12) was used to manage the data in a way that supported data analysis (QSR International Pty Ltd, 2018). The lead author directed the analysis. Coding by a second rater (VL) was undertaken to provide an opportunity to reflect on the coding approach, and to enhance the interpretive depth of the data (Braun & Clarke, 2021).

## RESULTS

### Participants

A total of 25 individual interviews were conducted with i) patients who had received the intervention (n=11), ii) secondary care clinicians responsible for delivering the intervention (n=8), and iii) GPs (n=6). Two of the eight secondary care clinicians were actively delivering the intervention (in an in-patient setting), and all GPs provided insights on future implementation. Six patients had received the intervention as a part of in-patient care and five patients provided perspectives on receiving the intervention from out-patient vascular, ulcer, or specialised CLTI clinics. Patient participants spoke about living with multiple conditions including diabetes, stroke, and amputation and receiving care from primary care (GPs, district nurses, diabetic nurses) secondary care (vascular teams), social care (carers), and family. Three patients were >70 years old but all patients were retired or unemployed and spoke about the impact of PAD on quality of life including managing pain, slow wound healing, poor sleep, reduced daily functioning, and limited mobility. Patient characteristics are shown in Table 1 and provider characteristics are detailed Table 2.

Of the patient participants contacted by the researcher (CL), 42% agreed to take part in an interview (fifteen people declined) and 82% of staff agreed to take part (1 person declined participation and 4 people did not respond to the invitation). Reasons for non-participation for patients included being unable to remember the intervention (n=5), deteriorating health (n=7), just home from an in-patient admission (n=1), being admitted for angioplasty (n=1) and getting back on with life (n=1). The reason provided by providers for declining participation was moving to a new role. Secondary care clinicians and GPs spoke about all three elements of the intervention and patients provided insights on the education leaflet and GP action letter. The descriptive themes were mapped to implementation theory using the CFIR, to explore connections between the themes, and to compare findings across participant groups. Four of the five overarching CFIR domains are represented across participant groups: intervention characteristics, outer setting, inner setting, and characteristics of individuals. The mapping of themes across CFIR domains and participant groups is included in ODS 2.

For intervention characteristics, all participant groups spoke about the relative advantage, adaptability, and design, quality, and packaging of the education leaflet, as well as the complexity in implementing the GP action letter. For outer setting, patient needs and resources (including barriers and facilitators to meet needs) were highlighted, with all participant groups explaining that a delayed referral to secondary care is a barrier to implementing the intervention. For inner setting, structural characteristics and networks and communications were identified by all groups as influences on implementation. For structural characteristics, participants explained that the ability to follow the advice in the education leaflet is often determined by the availability of supervised exercise programmes, difficulty ordering medication, and relying on carers for food shopping. For networks and communications, participants spoke about the need to work as a three-way team, and to have ongoing access to the specialist vascular team. For characteristics of individuals, staff values, attitude, knowledge, and skills to provide the intervention and patient readiness to take on the advice of the intervention were highlighted across participant groups. There was a particular emphasis on the individual stage of change sub-domain, with participants highlighting a need to engage patients in their care and to promote shared decision making. The patient-healthcare provider relationship was identified as a central influence relating to individual identification within the organisation, that is, how patients and staff perceive the organisation and their relationship with that organisation).

A final coding frame for patients, secondary care clinicians and GPs was developed following interpretive analysis. Because of space limitations, a summary of each theme is provided, and the subthemes are not elaborated in this article; the full coding framework is included in ODS 3. Four overarching themes were identified: *the potential to make a difference, a solution to address the gap in no man’s land, prioritising and making it happen* and *personalised information and supportive conversations for taking on the advice*. Similarities and differences between participant groups are presented where they arise.

### Theme 1: The potential to make a difference

GPs and secondary care clinicians felt that the LEGS intervention provides an opportunity to meet patient need and to support implementation of NICE guidance and spoke optimistically about using the intervention to improve patient outcomes and to standardise a minimum quality of care.

> *…it standardises the quality of care and standardisation is a big problem that we have in vascular surgery because guidance is so vague. So, I think it raises awareness and standardises that at least we ensure the patient gets a minimum of care* (QR15, secondary care clinician, UHL).

GPs and secondary care clinicians went on to explain how the intervention can be used to inform, empower, and support relationship building with patients.

> *I’ve gone through the leaflet with them… and knowing that the patient has understood I will spend a couple of minutes going through the leaflet to make sure. If they’ve got any questions I will try and take the opportunity to build my relationship with the patient so that they do understand* (QR08, secondary care clinician, UHL).

To further improve quality of care, providers identified the intervention as a way to support shared working across primary and secondary care, promoting a team approach and providing an opportunity for collaborative working. One GP explained how the ‘*patient is in the middle’* (QR23) and a secondary care clinician felt that ‘*we don’t sing off the same hymn sheet’* (QR08). One GP explained that w*e don’t ever see secondary care specialists* and a secondary care clinician explained *there’s always this assumption that the GP will sort it out* (QR13). It was acknowledged that shared working provides an opportunity to understand each other’s roles, to manage expectations and to learn from one another. Providers went on to report patient benefits in terms of access to specialist clinicians and working as a multi-disciplinary team, *I’ve always found that patients who have the opportunity to talk to multiple members of the medical team rather than a doctor or a nurse come out with better outcomes* (QR08, secondary care clinician, UHL).

Patients also spoke about the benefit of joined-up working, with one person stating;

> *…you attend lots of different clinics, you’ve got different people involved in your healthcare. It’s not just one thing and everything’s linked… it’s what they call whole person care. It’s not just your blood pressure, it’s also the vascular team, it’s the podiatrist, it’s the diabetes specialist* (QR07, out-patient clinic, GSTT).

### Theme 2: A solution to address the gap in no man’s land

All participant groups spoke about a need to target patients earlier and identified a service provision gap in preventative medicine and early diagnosis. One secondary care clinician stated;

> *I think, almost targeting people earlier on, so claudicants would be really good… claudicants at the very first stage of their peripheral arterial disease, they’re not really managed in secondary care, but they’re also not really managed in primary care… this gap where they’re in no man’s land and they almost wait until it’s severe enough to managed into secondary care* (QR18, secondary care clinician, UHL).

Patients explained how it took time to be referred to secondary care, and in one person’s situation, it became an emergency;

> *…my doctor prescribed pills and some cream for my foot but they didn’t do anything. And then it was only when I went back to podiatry after three months of constant pain, it was a case of just wait and see how things went… I went one day in the November, and they said you need to have your toe off almost immediately… they sent me to [hospital] and [hospital] say ‘your toe needs to come off and it needs to come off in 20 minutes time’* (QR04, In-patient care, UHL).

Difficulties with patients presenting late, high secondary care referral thresholds resulting in delayed referrals to secondary care and a wait for secondary care to make the diagnosis, alongside lack of support for primary care to commence diagnostics in the community were also listed as barriers to accessing timely care. Some secondary care clinicians felt that the care of those with IC should be managed in primary care.

> *I think even the way that if the GP could start the patient, the first medical therapies, like, clopidogrel and statins, if he also could start the patient on exercise therapies. So, it should be implemented at the same kind of wave of the first medical therapy*. (QR19, secondary care clinician, UHL).

GPs offered their support and made a request to do more sooner. Some suggested using the intervention in general practice as part of a PAD prevention programme. They acknowledged the value of using the intervention to promote earlier conversations with patients, manage patient expectations, and support primary care to start best medical therapy. GPs also spoke about how being involved sooner, and completing the clinical checklist, could be used to support triage of referrals to secondary care. GPs noted that preventative medicine and health promotion is core to their practice, and that sharing information across services and with patients reinforces the message and supports implementation.

### Theme 3: Prioritising and making it happen

Secondary care clinicians spoke about the need to prioritise the intervention alongside limited resources and the burden of work. One secondary care clinician explained, *it shouldn’t be seen as an extra thing* (QR08). Individual levels of confidence, attitude, knowledge and experience of the LEGS bundle, or a similar checklist/patient education intervention influenced prioritisation with some staff identifying checklists as a quick and easy-to-use memory prompt. However, another reported that;

> *…people feel that it’s just a tick-box exercise rather than a safety mechanism, and therefore no engagement with it… [it’s] another piece of paper… I think part of the challenge is engagement with clinicians. Clinicians don’t like threats to their autonomy… doctors, surgeons in particular are reticent, or certainly some are reticent to engage with checklists because they feel that it is doubling down or they already know this or that, you know, this simplifies things* (QR13, secondary care clinician, UHL).

Secondary care clinicians spoke about the extra resources required to deliver the intervention; it takes time to build relationships and to have conversations with patients. One secondary care clinician stated that finding the time to talk would require extra staff, this in an already stretched environment with staff shortages and long waiting times.

> *That will take more time, to make it personalised, to make it meaningful… you could almost imagine that they have a consultation and then some of those patients might benefit from sitting down with one of the specialist nurses… you could almost benefit from a 20-minute appointment after the standard clinic appointment to go through all of that. But again, that’s a resource burden, that means that you’ve got to have someone extra in clinic* (QR13, secondary care clinician, UHL).

Secondary care clinicians felt that the intervention fits the in-patient environment neatly and could be used as a record of the out-patient clinic consultation. GPs felt that it could support the complexity of assessing patients remotely in the community. The need for training was recognised;

> *I think there is a huge lack in primary care education, and I think that goes to show by the starting people coming in are only on like a 40% of them are on Best Medical Therapy so I think a big thing to target would be primary care to inform them* (QR18, secondary care clinician, UHL).

However, GPs acknowledged that teams are not static and require ongoing access to training GPs went on to request that all primary care staff be involved and not just GPs, and that the primary care training be delivered by the vascular team.

### Theme 4: Personalised information and supportive conversations for taking on the advice

Providers and patients spoke about the delivery of the intervention, recognising the importance of offering personalised care and tailoring patient information to individual needs. Secondary care clinicians felt that the intervention should be adapted to identify realistic targets and individual goals, *or the advice rings hollow* (QR13), and to offer a different approach for each patient. Staff reported;

> *I think the hard thing as well is that with the LEGS bundle, sometimes you’re giving it to someone who’s just had an amputation or about to have an amputation so it’s just to be mindful you’re talking about things like exercise they look at you very disparagingly and that can be a bit tricky… I think it’s a really good checklist and bundle for claudicants and I think that’s where it really shines but I think for when you get to the stage of CLTI, I think those individuals often feel that they’re too far gone for you to talk to them about lifestyle modification and things like that* (QR18, secondary care clinician, UHL).

Patients also spoke about the importance of timing in terms of when they received the lifestyle information.

> *If I was a bit younger and I’d got more get-up-and-go, I would gladly have a leaflet and go through it and take up advice and what have you. But as I am at the moment, my health is far from good* (QR14, in-patient care, UHL).
>
> *…it is a bit too late in that respect so I can only do what I can do. I mean yesterday I was out in the garden doing some weeding but I have to use a seat to do it and the day before I was cutting the grass and I have a seat at the top of the garden and a seat down at the bottom of the garden and I’ll do maybe three stripes and then I have to have a little sit down just to get my leg going again* (QR05, in-patient care, UHL).

Patients explained that they like to receive information face-to-face and suggested that the education leaflet could be used as a supportive conversation guide. They emphasised the importance of the conversation that goes alongside the advice and information, and the relationship that they have with their health professional(s) in terms of asking questions and being able to talk about making changes like smoking cessation. One patient explained,

> *… the only thing I’ve got to do is drink and smoke and watch the tv – cause if I didn’t have those three things, I would be pulling my hair out… to be quite honest, I’m quite happy at the moment to be carrying on the way I am. It might sound silly to you, but I enjoy smoking. I love a drink, I like my telly… if you’re brow-beaten into packing up when you don’t really want to, all this, it can have a great effect on people* (QR14, in-patient care, UHL).

GPs and secondary care clinicians acknowledged the need to support patient preference, to involve patients in the conversation and to share decision making.

> *…you have to strike a balance, and therefore taking the patient’s preferences into account is important. We assume that patients want to live the longest life they can, I’m not sure that’s always correct. Some patients would rather accept that they live a slightly shorter life but with the benefit of enjoying certain things that we all know are less healthy* (QR13, secondary care clinician, UHL).

It was acknowledged that everyone knows they need to stop smoking and it is easy to bombard people with information. Solutions such as motivational interviewing techniques were identified by one GP as a way to support engagement conversations. Secondary care clinicians felt that providing information on the benefits of following the advice versus taking pleasure away might help. For others, frank conversations that explain the risks help;

> *I think honesty is the most important aspect of this and bringing people in front of the data, highlighting the data, and actually not sugar coating anything. We know, for example, that if you have critical ischemia, so severe peripheral arterial disease, if your prognosis is worse, your five-year survival is worse than some kinds of cancer. So, if you tell someone that unless you actually take this intervention seriously, your likelihood of not being around in five years is very high. I know it sounds bleak, but it may be the wake-up call that someone needs. Or explaining that from the moment you have severe arterial peripheral disease, the likelihood of you losing your leg and being wheelchair bound may be an important thing to highlight as opposed to speaking in a circumspect way. I think just being honest and explaining the risks may be a way of engaging patients*. (QR15, secondary care clinician, UHL).

A patient explained how the honest frank information helped;

> *The doctor actually threatened me. He said to me, if I’m going to do this operation, you got to stop smoking. So, I did. That helped, or I could lose my leg. I thought it was brilliant the way [the doctor] come out with it and I’ve never stopped thanking him for it. It works* (QR06, out-patient care, GSTT).

## Discussion

The aim of the study was to provide an understanding of the acceptability of the LEGS (LEaflet Gp letter Structured checklist) intervention, a complex intervention in the form of a healthcare bundle, designed to improve the care of patients with PAD in the NHS. Our inductive approach to analysis found four overarching themes: i) *the potential to make a difference, ii) a solution to address the gap in no man’s land, iii) prioritising and making it happen* and *iv) personalised information and supportive conversations for taking on the advice*. We used the Consolidated Framework of Implementation Research (CFIR) to guide our analysis, and all five CFIR domains are represented (Damschroder et al., 2009).

All participants spoke positively about using the intervention to make a difference to patient care and as an opportunity to meet patient needs. Moreover, a way to enhance the quality of PAD care through improved working practices. This promotes a streamlined patient pathway from primary care to secondary care and to shared care. There was a consensus of optimism in participant accounts about the potential to make a difference. The findings confirm that the period between diagnosis of early-stage PAD (e.g., claudication) and subsequent potential diagnosis of advanced PAD with severe symptoms, is a time when patients do not receive any help, education, support, or appropriate medication to address their condition (Saratzis et al., 2019). As such, the benefits of the LEGS intervention were identified in terms of meeting the needs of this subgroup of patients, and to address the gap in care. GPs were optimistic that they can deliver the LEGS intervention with training and ongoing sustained communication with secondary care. The findings highlight the need to consider training requirements and systems for maintaining open primary-secondary care communication to support implementation. For example, education on tailoring the intervention for patients at both early and late stages of PAD, that can be used across primary and secondary care settings, similar to the model used with other chronic conditions (Smith et al., 2017).

While the three elements of the LEGS intervention have been designed to facilitate interaction between secondary and primary care following diagnosis of PAD, the intervention focus until now, has been on secondary care services, that is, hospital admission or out-patient clinic. However, our findings indicate that the intervention can be used to address the gap in service provision for patients who do not yet meet the referral criteria for secondary care services. We therefore propose to extend the use of the LEGS intervention across the primary-secondary care interface. Our findings also confirm the need for individualised patient care, supportive communication and shared decision making in order to take on the advice in a more effective way (Barry & Edgman-Levitan, 2012). The finalised LEGS intervention should therefore promote effective conversation between patient and healthcare staff, both in primary and secondary care settings. The open approach to changing lifestyle and behaviour could be built on techniques like motivational interviewing, which is already commonly used in primary care practice (Rubak et al., 2005). Lastly, our findings confirm that healthcare providers are optimistic and positive about the introduction of an intervention which will help them to deliver more consistent and guideline-based PAD treatment. The challenge now is to take this enthusiasm together with the lessons learned from this qualitative research to adapt the LEGS intervention before wider testing and adoption in the NHS. The main changes will relate to usability in primary care after diagnosis of early-stage PAD, facilitation of effective communication not only between primary and secondary care staff, but also patients and staff, as well as focussing on specific needs of patients by prioritising areas to focus on in an incremental or stepwise manner.

## Strengths and limitations

While the paper adds new knowledge on the development and implementation of a community and hospital intervention for Peripheral Arterial Disease, it is important to note that the findings are specific to the CHABLIS study. It is possible, however, to enhance transferability by describing the research context and assumptions, and by making connections between the analysis of participants accounts and claims in the extant literature. A strength is the focus on rigor and triangulation of participant perspectives. We also acknowledged the complexity of intervention implementation across the secondary-primary care interface and therefore used a systems research perspective for addressing implementation at the outset of the research. We used implementation theory to guide analysis and to provide a layered coding approach, to ensure that intervention and implementation recommendations are novel, grounded in the data, and useful (Damschroder et al., 2009).

While the anonymity of telephone use can allow participants to disclose sensitive information, telephone interviews have received criticism for compromising interviewer/participant rapport and interaction, and for limiting contextual data due to the absence of face-to-face contact and visual cues. However, this method of data collection is convenient, in that it is flexible (in terms of time and location), accessible (i.e., remote research conducted during the COVID-19 pandemic), and allows for a wide reach (e.g., accessing a diverse population from both urban and rural areas) (Irvine, 2011).

## Conclusions

The LEGS intervention can be tailored for use at early and late stages of PAD, can be provided across primary and secondary care settings, and provides an opportunity to promote shared working across the primary-secondary care interface. These findings are important to the specifics of the CHABLIS study but can also go some way in informing the wider learning for other interventions that are implemented across the primary-secondary care interface.

## Supporting information

ODS CFIR Domains across participant groups

ODS Interview Schedules

Tables 1 and 2 - participant characteristics

## Data Availability

Available freely after contact with chief investigator.

## Declarations

### Ethics approval and consent to participate

The research was reviewed and approved by the NHS Wales Research Ethics Committee (REC), reference number20/WA/0319 and the NHS Health Research Authority (HRA). It was funded by the National Institute for Health Research NIHR Research for Patient Benefit (RfPB) programme (funder’s reference: NIHR202008). The study was prospectively registered, and the full protocol has been published online (reference: SRCTN13202085, available at: https://isrctn.com/ISRCTN13202085).

### Consent for publication

Consent for publication has been obtained from all research participants.

### Availability of data and materials

This study uses data (containing potentially identifying and/or sensitive information) collected from a small group of staff participants and a vulnerable patient population, and involves indirect identifiers (such as sex, ethnicity, location, etc.) that may risk the identification of study participants. Sharing data outside of the anonymised excerpts and quotations included in the paper will violate the agreement to which the participants consented.

### Authors’ contributions

CL contributed to the qualitative study design, coordinated the study, led data collection and analysis, and drafted the manuscript. AS is principal investigator to the CHABLIS study, acquired funding, participated in the study design, contributed to data collection, and drafted the manuscript. PS contributed to data collection and reviewed the manuscript. RB and BB drafted the manuscript. EW contributed to data collection and reviewed the manuscript. VL designed the qualitative study, provided oversight on study coordination, data collection and analysis, and drafted the manuscript. All authors read and approved the final manuscript.

## Acknowledgements

We would like to thank all patients, secondary care staff and GPs who participated in this study and who generously gave their time and honest thoughts. We would like to thank the local research teams for their support in recruiting to this qualitative study and we are also grateful to the NIHR Clinical Research Network Research Delivery Managers that helped to recruit GP participants.

This article presents independent research funded by NIHR Research for Patient Benefit (RfPB) Programme (NIHR202008). The views expressed are those of the author(s) and not necessarily those of the NIHR or the Department of Health and Social Care.

## Online data supplements

File 1: Interview schedule for patients and providers

File 2: Table of themes across participant groups

File 3: Full coding framework

## List of abbreviations

PAD: Peripheral Arterial Disease
IC: Intermittent Claudication
CLTI: Chronic Limb Threatening Ischaemia
GP: General Practitioner
CHABLIS: Community and Hospital cAre Bundle to improve the medical treatment of cLaudIcation and critical limb iSchaemia
LEGS: LEaflet Gp letter Structured checklist
UHL: University Hospitals of Leicester NHS Trust
GSTT: Guy’s and St Thomas’ NHS Foundation Trust
CFIR: The consolidated framework for implementation research
NHS: National Health Service
ODS: Online Data Supplement

