## Supplementary material for "Patient and provider perspectives on factors that influence the implementation of a community and hospital care bundle to improve the treatment of patients with peripheral arterial disease in primary and secondary care": ODS CFIR Domains across participant groups

| CFIR Domains and the LEGS bundle | Patients | Secondary care clinicians | GPs |
| --- | --- | --- | --- |
| **THE LEGS BUNDLE** | | | |
| Element 1: Clinical checklist |  | X | X |
| Element 2: Patient and carer education leaflet | X | X | X |
| Element 3: GP action letter | X | X | X |
| **CFIR DOMAINS** | | | |
| **Domain 1: INTERVENTION CHARACTERISTICS** | **X** | **X** | **X** |
| **1.1 Intervention source:** Perception about whether the intervention is externally or internally developed. | | | |
| The whole bundle |  | X | X |
| I sat down with the developers, and they gave me the background |  | X |  |
| Perception about how the intervention was developed |  |  | X |
| Involving stakeholders in the development process will promote uptake |  |  | X |
| **1.2 Evidence strength and quality:** Perceptions of the quality and validity of evidence supporting the belief that the intervention will have desired outcomes. | | | |
| Clinical checklist |  | X |  |
| Built on existing NICE guidance |  | X |  |
| The guidance is vague |  | X |  |
| The guidance is clear, but we don’t follow it |  | X |  |
| **1.3 Relative advantage (or not):** Perception of the advantage of implementing the intervention versus an alternative solution. | | | |
| The whole bundle |  | X | X |
| Improves care and outcomes |  | X |  |
| Standardises a minimum quality of care |  | X |  |
| Allows us to make a difference to patients |  | X |  |
| Joins up primary and secondary care working |  |  | X |
| Improves awareness of NICE guidance in primary & secondary care |  | X | X |
| Improves awareness and understanding of PAD |  | X | X |
| Clinical checklist |  | X |  |
| Builds your relationship with the patient |  | X |  |
| Improves medication compliance |  | X |  |
| Patient and carer education leaflet | X | X | X |
| Patients are empowered by being informed |  | X |  |
| I don’t want any more leaflets | X |  |  |
| District nurses can answer my questions | X |  |  |
| A conversation guide for primary care |  |  | X |
| Existing relationships to reinforce the message |  |  | X |
| GP action letter |  | X | X |
| A standardised letter with clear information |  | X |  |
| Useful for non-medical led community vascular clinics |  |  | X |
| Repeat GP action letter is a duplicate, adds to the workload and sets the wrong tone |  |  | X |
| It gives the impression that secondary care do not trust GPs |  |  | X |
| **1.4 Adaptability:** The degree to which an intervention can be adapted, tailored, refined, or reinvented to meet local needs. | | | |
| Clinical checklist |  | X |  |
| Adapting to the environment |  | X |  |
| Fits the in-patient environment neatly |  | X |  |
| Incorporate into a universal admission checklist |  | X |  |
| Limited access to blood pressure monitoring or cholesterol tests in out-patient clinic |  | X |  |
| Promote checklist in primary care to be used alongside secondary care referral letter |  | X |  |
| Patient and carer education leaflet | X | X | X |
| Offer personalised care and tailor patient education to individual needs |  | X | X |
| A different approach for each type of patient |  |  | X |
| Different information for each patient |  |  | X |
| Existing info most likely to suit those with intermittent claudication |  |  | X |
| Offer realistic targets or the advice rings hollow |  | X |  |
| Consider co-exiting morbidities |  |  | X |
| Too much information | X |  |  |
| Change information layout to provide summary at outset |  | X |  |
| Missing information | X |  | X |
| Understanding what causes PAD and preventing further blockages | X |  | X |
| Managing the psychological impact of PAD | X |  |  |
| Maintaining movement and physiotherapy input | X |  |  |
| Impact on work, social and family life | X |  |  |
| Acknowledgement of reliance and impact on family | X |  |  |
| Carer and family support | X |  |  |
| Managing future risks and when to seek support |  | X | X |
| **1.5 Trialability:** The ability to test the intervention on a small scale in the organisation, and to be able to reverse course (undo implementation) if warranted. | | | |
| Clinical checklist | X | X |  |
| A quick quality check |  | X |  |
| Patient and carer education leaflet | X | X |  |
| Leaflet is a resource for referring back | X | X |  |
| **1.6 Complexity:**  Perceived difficulty of implementation, reflected by duration, scope, radicalness, disruptiveness, centrality, and intricacy and number of steps required to implement. | | | |
| Clinical checklist |  | X | X |
| Simplicity |  | X |  |
| Improvements that make it even simpler |  | X |  |
| It needs to be a bit more, no and why |  | X | X |
| GP action letter | X | X | X |
| A template for new staff |  | X |  |
| A time-consuming manual coding process with potential for error |  |  | X |
| Waiting times for primary care/test appointments | X |  | X |
| Difficult to assess patients remotely |  |  | X |
| Lack of continuity |  |  | X |
| 60 calls and 40 letters a day |  |  | X |
| Up to three months to receive hospital letters |  |  | X |
| Patients to hand deliver letter to GP surgery |  |  | X |
| **1.7 Design quality and packaging:** Perceived excellence in how the intervention is bundled, presented, and assembled. | | | |
| Clinical checklist |  | X |  |
| Working across two different systems - Paper and electronic |  | X |  |
| Paper checklists tend to get missed |  | X |  |
| Electronic option required |  | X |  |
| Patient and carer education leaflet | X | X | X |
| Accessibility | X |  |  |
| Stored in a safe place | X |  |  |
| Lost the leaflet | X |  |  |
| Would be improved if available by email | X |  |  |
| Presentation & format | X |  |  |
| Paper leaflet |  | X |  |
| Larger print has been requested |  | X |  |
| Language can be hard to understand | X |  | X |
| Write for a lower reading age and provide additional information or additional resources for people who are able to access more detailed information |  |  | X |
| Keep it simple, give less information, in different formats and in a way that is understandable |  |  | X |
| Avoid didactic language and reframe the approach |  |  | X |
| There’s a lot of ‘do nots’ |  |  | X |
| Ensure accuracy of information |  |  | X |
| Confusing dietary advice |  |  | X |
| Provide more infographics and pictures | X | X | X |
| Include diversity in visuals (e.g., pictures of leg ulcers/skin changes in different skin colours) |  | X |  |
| Add web links |  |  | X |
| Could be supported by existing NHS software like QRX, direct information to patient phones |  |  | X |
| Offer an audio and video version | X | X | X |
| Patient information to be offered as a group intervention | X | X |  |
| Opportunity for group to be peer-led |  | X |  |
| GP action letter |  |  | X |
| Same information and same system across primary and secondary care |  |  | X |
| Include information on the discussion around contraindications and side effects of any newly prescribed medication with patient |  |  | X |
| Any contraindications primary care should be aware of, useful for clarification |  |  | X |
| Offered and accepted/declined smoking cessation referral |  |  | X |
| Provide a clear plan for if the situation deteriorates |  |  | X |
| Copy of letter to patient – shared message – reinforcement |  |  | X |
| **1.8 Cost:** Costs of the intervention and costs associated with implementing that intervention including investment, supply, and opportunity costs. |  |  | X |
| It is not commissioned or financially viable |  |  | X |
| Primary care time |  |  | X |
| Maintaining equipment |  |  | X |
| **Domain 2: OUTER SETTING** | X | X | X |
| **2.1 Patient needs and resources:** The extent to which patient needs, as well as barriers and facilitators to meet those needs are known and prioritised. | | | |
| The whole bundle | X | X | X |
| Working as part of a whole team |  |  | X |
| Promoting joined up working across primary and secondary care |  | X | X |
| Enabling specialist care from different people |  |  | X |
| Expectations and understanding of each other’s roles |  |  | X |
| Timely access back into secondary care if needed |  |  | X |
| Secondary care oversight |  |  | X |
| Accessible referral pathway | X |  | X |
| We can’t make a referral anyway |  |  | X |
| Secondary care referral thresholds too high |  |  | X |
| You don’t fit the criteria so wait until it gets worse |  |  | X |
| Delayed referral to secondary care | X | X | X |
| Presenting too late |  | X |  |
| Can I do some of the diagnostics in the community? |  |  | X |
| Time wastage |  | X | X |
| A geographical consideration | X |  | X |
| Locality of secondary care services | X |  | X |
| Transport needs | X |  | X |
| Clinical checklist |  | X | X |
| Use in primary care to start best medical therapy |  | X |  |
| Would promote earlier conversations with patients in primary care |  | X |  |
| Use at point of admission and discharge for in-patients |  | X |  |
| Adapt for primary care to support referral |  |  | X |
| Patient and carer education leaflet | X | X | X |
| Patient education provided too late | X | X |  |
| Too far gone to talk about lifestyle modification |  | X |  |
| Level of functioning & mobility - I can only do what I can do | X |  |  |
| It is crucial whenever it happens |  | X |  |
| Introduced sooner in primary care as part of a PAD prevention programme |  |  | X |
| Informing patients to reduce the chance of presenting late/in crisis |  |  | X |
| Managing patient expectations |  |  | X |
| **2.2 Cosmopolitanism:** The degree to which an organisation is networked with other external organisations. | | | |
| The whole bundle |  | X |  |
| Shared primary & secondary care working |  | X |  |
| Patient and carer education leaflet |  |  | X |
| Copy of leaflet to primary care to reiterate message |  |  | X |
| GP action letter |  | X |  |
| Clear open communication across community and hospital services |  | X |  |
| Joined up clinical record systems |  | X |  |
| We don’t sing off the same hymn sheet |  | X |  |
| **2.3 External policies and incentives:** A broad construct that includes external strategies to spread interventions including policy and regulations (governmental or other central entity), external mandates, recommendations and guidelines, pay-for-performance, collaboratives, and public or benchmark reporting. | | | |
| The whole bundle |  | X |  |
| Additional funding |  | X |  |
| **Domain 3: INNER SETTING** | X | X | X |
| **3.1 Structural characteristics:** The social architecture, age, maturity, and size of an organisation. | | | |
| The whole bundle |  | X | X |
| Impact of staff shortages |  | X |  |
| Reduced staffing due to covid |  | X |  |
| Involving the multi-disciplinary team |  |  | X |
| GP action letter |  | X | X |
| Time delay between writing and getting to primary care |  | X | X |
| Patient and carer education leaflet | X | X | X |
| Being able to follow the advice | X | X |  |
| Difficulty ordering repeat medication | X |  |  |
| Relying on others for food shopping | X |  |  |
| Availability of supervised exercise |  | X |  |
| It’s easy to bombard somebody with information |  |  | X |
| **3.2 Networks and communications:**  The nature and quality of webs of social networks and the nature and quality of formal and informal communications within an organisation. | | | |
| The whole bundle | X | X | X |
| A team approach |  | X |  |
| Building relationships |  |  | X |
| Clear communication |  |  | X |
| The patient is in the middle |  |  | X |
| Involve all multi-disciplinary professionals (including pharmacists, nurses, physios) |  | X |  |
| Access to the vascular team or not | X |  | X |
| Advice and information | X |  | X |
| **3.3 Culture:** Norms, values, and basic assumptions of a given organisation. | | | |
| The whole bundle |  | X |  |
| Target patients earlier |  | X |  |
| The gap in no man’s land |  | X |  |
| **3.4 Implementation climate - tension for change:** The degree to which stakeholders perceive the current situation as intolerable or needing change. | | | |
| The whole bundle |  | X | X |
| Primary care needs to do more sooner |  | X |  |
| Secondary care referral threshold is too high |  |  | X |
| **3.5 Implementation climate - Compatibility:** The degree of tangible fit between meaning and values attached to the intervention by involved individuals, how those align with individuals’ own norms, values, and perceived risks and needs, and how the intervention fits with existing workflows and systems. | | | |
| The whole bundle |  |  | X |
| Existing systems in place for primary care medication management and monitoring |  |  | X |
| Provide 28 days medication to allow for delay in seeing GP |  |  | X |
| Preventative medicine and health promotion is core to primary care practice |  |  | X |
| Ability to offer smoking cessation support |  |  | X |
| Local variation in set-up of smoking cessation, dietary advice, exercise therapy |  |  | X |
| No formal set-up in primary care to pick it up |  |  | X |
| GP relationship with patients |  |  |  |
| Not commissioned |  |  |  |
| Secondary care to initiate smoking cessation referral |  |  | X |
| National protocol for weight reduction |  |  | X |
| Existing relationships with Diabetes nurses/clinics |  |  | X |
| **3.6 Implementation climate - Relative priority:** Individuals’ shared perception of the importance of the implementation within the organisation. | | | |
| Clinical checklist |  | X |  |
| Take the opportunity |  | X |  |
| It shouldn’t be seen as an extra thing |  | X |  |
| A threat to autonomy |  | X |  |
| Checklists are tick-box exercises rather than safety mechanisms |  | X |  |
| **3.7 Implementation climate - Learning climate** | | | |
| The whole bundle |  |  | X |
| Two-way learning |  |  | X |
| Secondary care can learn from this too |  |  | X |
| Holistic picture of medication profile, side effects, competing medication |  |  | X |
| Teams are not static so require ongoing access to training |  | X | X |
| **3.8 Readiness for implementation – leadership engagement:** Commitment, involvement, and accountability of leaders and managers with the implementation. | | | |
| The whole package |  | X |  |
| Provide a named leader |  | X |  |
| External oversight is preferable |  | X |  |
| **3.9 Readiness for implementation – available resources:** The level of resources dedicated for implementation and on-going operations including money, training, education, physical space, and time. | | | |
| Clinical checklist |  | X | X |
| Checklists are quick |  | X |  |
| Managing it alongside the burden of work |  | X |  |
| The assumption that the GP will sort it – putting it back into GP land |  | X | X |
| Lack of access to testing equipment in community |  |  | X |
| Limited community workforce (i.e., reduced number of district nurses) capacity for conducting ABPIs |  |  | X |
| Secondary care are gatekeepers for ABPIs and duplexes |  |  | X |
| Waiting for secondary care to make diagnosis |  |  | X |
| Patient and carer education leaflet |  | X |  |
| Time to talk requires extra staff |  | X |  |
| GP action letter |  | X | X |
| Time burden and impact of dictating long clinic letters |  | X |  |
| Joining it up - We’ve done our bit and now pass it on to the GP |  |  | X |
| Routine practice or adding to the workload |  |  | X |
| Feels top down and with didactic language |  |  | X |
| Keep the GP onside and change phraseology |  |  | X |
| **3.10 Readiness for implementation - access to information and knowledge:** Ease of access to digestible information and knowledge about the intervention and how to incorporate it into work tasks. | | | |
| The whole bundle |  | X |  |
| Training on the purpose of the intervention |  | X |  |
| Written information for those unable to attend training |  | X |  |
| Prioritise training and supervision for junior staff – the people who do the day-to-day work |  | X |  |
| Consider involving medical students |  | X |  |
| Highlight the link between PAD and cardiovascular death |  | X |  |
| Education on managing CLTI and Non-CLTI patients in primary care |  | X |  |
| Knowing the risk factors for CLTI and how to make a referral to secondary care |  | X |  |
| Taking an ABPI in primary care |  | X | X |
| Support the triage for secondary care |  |  | X |
| We’re not commissioned to do it |  |  | X |
| Visibility of vascular teams at primary care conferences |  | X |  |
| Learning on the job |  | X | X |
| Informal training to colleagues i.e., when seeing a medical outlier |  | X |  |
| Use available supervision/debriefing opportunities |  |  | X |
| Webinar-ed out |  | X |  |
| Training delivered by vascular team to primary care |  |  | X |
| Collaborative working and building relationships |  |  | X |
| We don’t ever see secondary care specialists |  |  | X |
| Involve all primary care staff and not just GPs |  |  | X |
| Learning gap in exercise and lifestyle modification |  |  | X |
| **DOMAIN 4: CHARACTERISTICS OF INDIVIDUALS** |  |  |  |
| The whole bundle | X | X | X |
| Taking on the advice | X | X | X |
| Familiarity and understanding of PAD |  | X | X |
| **4.1 Knowledge and beliefs about the intervention** | | | |
| Clinical checklist |  | X | X |
| Prioritising the checklist (or not) |  | X |  |
| Previous experience of using checklists and identifying benefits |  | X |  |
| Checklists are a useful memory prompt |  | X |  |
| Patient and carer education leaflet | X |  |  |
| Targets that are impossible to reach | X |  |  |
| **4.2 Self-efficacy** | | | |
| The whole bundle |  | X |  |
| Confidence to implement guidelines |  | X |  |
| **4.3 Individual stage of change** | | | |
| Clinical checklist |  | X |  |
| Put myself forward to find out more |  | X |  |
| Patient and carer education leaflet | X | X | X |
| If I was younger and I had more get-up-and-go | X |  |  |
| It’s down to willpower at the end of the day (I like smoking, I like drinking, I like the telly) – patient engagement & stage of change – resistance from patients | X | X | X |
| The only thing left is something nice to eat | X |  |  |
| Resistance from patients |  | X |  |
| Secondary care doctors tell people to do things |  |  | X |
| Everybody knows they need to stop smoking |  |  | X |
| **4.4 Individual identification with organisation:** how individuals perceive the organization and their relationship and degree of commitment with that organisation. | | | |
| The whole bundle |  | X |  |
| Joined up working with primary care |  | X |  |
| Patient and carer education leaflet | X | X | X |
| The patient – health professional relationship | X | X |  |
| It takes time to build the relationship and have the conversation |  | X |  |
| Clear open communication | X |  |  |
| Frank conversations for some | X | X |  |
| Be honest and explain the risks |  | X |  |
| Information on the benefits versus taking pleasure away |  | X |  |
| Striking a balance to optimise people’s quality of life. You can’t take everything away. |  | X |  |
| Patient preferences and shared decision making |  | X |  |
| Online tools to support the clinical conversation |  | X |  |
| Language and approach used to engage people |  |  | X |
| Consider use of motivational interviewing techniques in secondary care |  |  | X |
| GP action letter |  |  | X |
| Partnership working |  |  | X |
| Avoid the extra step and order the test in secondary care |  |  | X |
| **4.5 Other personal attributes:** other personal traits such as tolerance of ambiguity, intellectual ability, motivation, values, competence, capacity, and learning style. |  |  |  |
| Clinical checklist |  | X |  |
| Team members who prioritise PAD |  | X |  |
| Patient and carer education leaflet | X |  |  |
| Living with other health conditions alongside (arthritis, diabetes) | X |  |  |
| Managing the pain | X |  |  |
| Medication side-effects | X |  |  |
| **DOMAIN 5: PROCESS** |  |  |  |
| **5.1 Engaging:** Attracting and involving appropriate individuals in the implementation and use of the intervention through a combined strategy of social marketing, education, role modelling, training, and other  similar activities. | | | |
| The whole bundle |  | X |  |
| Championing the cause |  | X |  |
| Doing the bare minimum and going home on time |  | X |  |
| Using research to demonstrate a difference |  | X |  |
| Involving stakeholders at an early stage |  | X |  |
| Clinical checklist |  | X |  |
| Posters in clinic to remind staff |  | X |  |
| **5.2 Executing:** Carrying out or accomplishing the implementation according to plan. | | | |
| Patient and carer education leaflet | X |  |  |
| A direct contact number for the vascular team | X |  |  |
| Primary care capacity | X |  |  |
| My GP and practice nurses keep an eye on me | X |  |  |
| Sometimes it can take weeks to get them to answer a query over the phone | X |  |  |
| GP action letter | X |  |  |
| It can take weeks to get information on their system | X |  |  |
| It’s lots of different clinics and different people involved in your healthcare (whole person care) | X |  |  |

**CFIR matrix and distribution of descriptive themes**
