## Supplementary material for "Patient and provider perspectives on factors that influence the implementation of a community and hospital care bundle to improve the treatment of patients with peripheral arterial disease in primary and secondary care": ODS Interview Schedules

**Interview Topic Guide for Patients**

**Title of the study:** A **C**ommunity and **H**ospital c**A**re **B**undle to improve the medical treatment of c**L**aud**I**cation and critical limb i**S**chaemia. **(**The CHABLIS study)

**Welcome and introduction**

*[Seek consent to continue and to audio-record the interview (if applicable). Participant has to provide written informed consent (see separate form). Let them know that no personal identifiable data will be recorded and a participant number will be allocated to them.]*

The main aim of this study is to assess whether a dedicated intervention (a care bundle) can be used in the routine care of patients who present with symptomatic Peripheral Arterial Disease (PAD). The care bundle includes a number of checklists and leaflets for doctors and patients. The intervention is aimed at reducing the risk of major cardiovascular events (i.e. heart attacks and strokes), and preventing amputation, by making sure patients with PAD receive the appropriate treatment and management as recommended by national guidelines. We have called this intervention LEGS (the LEGS bundle), given that the aim is to save legs from amputation and improve vascular care.

The LEGS intervention:

ELEMENT 1: In-patient checklist/Outpatient Checklist

The doctors who see you will be given a checklist of the common things that patients with PAD should be offered in terms of medications and / or tests. All of these suggestions are already part of what is the standard NHS care for PAD. Using your medical notes, we will record your blood pressure, your medications (current), you medical history, previous surgeries, imaging results, your routine blood test results (cholesterol, lipid profile, full blood count), smoking status, and demographic information (age, height, weight, gender). This is standard information that the NHS already collects in order to be able to offer you treatment for your blocked arteries.

ELEMENT 2: 2^nd^ (follow-up) letter to the GP

ELEMENT 3: Patient & carers’ leaflet

You will be given a leaflet which will provide you with information about PAD and other things you can do to help reduce your risks.

You will have an appointment to see us again in six months to check your legs and your overall health. This is standard for someone with PAD. We will again record your blood pressure, your medications (current), your medical history, previous surgeries, imaging results, your routine blood test results (cholesterol, lipid profile, full blood count), smoking status, and demographic information (age, height, weight, gender). During that appointment we will ask you to fill in a questionnaire which assesses how well people are taking their prescribed medication, called the MARS questionnaire.

The interview will explore your experience of receiving the intervention. We will ask about your opinion of NHS care relating to your leg artery problems in greater detail. We will ask you how you have found the management of your care at hospital and your GP practice, what your thoughts are of the information leaflet, and if you think it has been of any benefit to you or not.

**Questions:**

1. How have you found the management of your care at hospital and your GP practice?

- the initial consultation between patient and healthcare professional after the diagnosis of peripheral arterial disease has been made
  - what should be covered in the initial consultation?
  - Anything missing from this consultation?
  - Are you able to ask questions?
- the frequency and duration of follow-up

2. What are your thoughts on the information leaflet?

- Is there enough information about peripheral arterial disease? Too much information? Is the information clear? Signposts for where to go for more information?
- Is there anything on the leaflet that you don’t understand or would like more information about?
- What would you like to see included in a leaflet designed to inform patients about peripheral arterial disease care and therapies?
- What about the length of the leaflet?

3. Do you think it has been of any benefit to you or not?
4. In your opinion, are there any factors that might influence someone’s ability to follow the guidance in the patient information leaflet? (i.e. barriers to taking medication, managing diet)

- Is there anything that could help? Solutions to any barriers for following the advice?

5. Any ideas for improving any of the elements of the intervention?

ELEMENT 1: Inpatient/outpatient checklist (& patient consultation)

ELEMENT 2: 2^nd^ (follow-up) letter to the GP (& patient consultation)

ELEMENT 3: Patient & carers’ leaflet

6. What do you need for your health to improve? (wider experiences and perceived needs)

**Anything not covered?** Is there anything that we haven’t covered in the interview that you think we should know or think about?

**Closing and thanks -** check that the participant is still happy for you to use all the information provided and offer the possibility to erase sections of the recording.

Thank them for their time and contribution.

**Interview Topic Guide for Secondary Care Clinicians & GPs**

**Title of the study:** A **C**ommunity and **H**ospital c**A**re **B**undle to improve the medical treatment of c**L**aud**I**cation and critical limb i**S**chaemia. **(**The CHABLIS study)

**Welcome and introduction**

*[Seek consent to continue and to audio-record the interview (if applicable). Participant has to provide written informed consent (see separate form). Let them know that no personal identifiable data will be recorded and a participant number will be allocated to them.]*

The main aim of this study is to assess whether a dedicated intervention (a care bundle) can be used in the routine care of patients who present with symptomatic Peripheral Arterial Disease (PAD). The care bundle includes a number of checklists and leaflets for doctors and patients. The intervention is aimed at reducing the risk of major cardiovascular events (i.e. heart attacks and strokes), and preventing amputation, by making sure patients with PAD receive the appropriate treatment and management as recommended by national guidelines. We have called this intervention LEGS (the LEGS bundle), given that the aim is to save legs from amputation and improve vascular care.

ELEMENT 1: In-patient checklist/Outpatient Checklist

ELEMENT 2: 2^nd^ (follow-up) letter to the GP

ELEMENT 3: Patient & carers’ leaflet

The interview will explore your experiences/perspectives of delivering the intervention. This will help us to evaluate the ability/feasibility of staff in delivering the intervention, overall impression, validity, usability, acceptability and engagement with the intervention. It will include barriers to engagement or delivery and ideas for improving the intervention.

**This is a GUIDE for topics. Further topics will and can be explored if necessary.**

The purpose of the study is to find out about staff views and experiences of implementing the LEGS intervention as a part of routine clinical care. This provides us with information on factors that facilitate or hinder implementation and on possible solutions to overcome any identified barriers.

Key objectives of the interview session: i) to identify barriers to and facilitators of each of the key intervention targets; ii) identify issues around delivery, training needs and competencies.

***We would like to ask you about what helps and what hinders success in implementing the LEGS intervention as a part of routine clinical care.***

•	To get started, we would like to ask about your views on existing NICE guidance and factors that might influence implementation of NICE guidance on cardiovascular risk reduction amongst patients with peripheral arterial disease. Please comment on: i) prescribing, ii) awareness of NICE guidance by primary and secondary care, iii) adherence by patients (when they have been prescribed their medication), iv) follow-up practices in primary and secondary care.

•	What do you think about using the LEGS intervention with this group of patients? Are there any benefits? Or challenges?

Understanding implementation

Aims:	1) To explore what participants think helps or hinders implementation.

2) To explore what participants’ identify as potential solutions to implementation barriers.

**What helps implementation?**

•	What factors do you perceive might support implementing the LEGS intervention in routine clinical practice?

•	What helps/would help you (and your colleague) to provide the intervention a part of routine clinical care?

•	What are your views on the LEGS intervention and the acceptability and feasibility of delivering the intervention?

•	Do you feel that you have the knowledge and skills to deliver this intervention?

•	What implementation strategies might help? Training?

**What prevents or hinders implementation?**

•	What prevents or hinders success in implementing the LEGS intervention as a part of routine clinical care?

•	Are there/or might there be challenges around implementation?

•	What would further support you to implement the intervention?

Prompts / Continuation Questions:

•	What resources would better equip you to implement the intervention?

•	What would you say is the ideal environment to implement the LEGS intervention in?

**Implementation solutions**

•	What solutions would you recommend to address these identified barriers?

•	Any other influences on implementation? COVID? Other clinical priorities?

•	Any changes needed to the intervention? Ideas for improving the intervention?

(Can you advise us on the 3 most important things you would like to see covered in a patient leaflet for patients with peripheral arterial disease? Can you please advise us on the 3 most important things you would like to see covered in a checklist which supports the delivery of peripheral arterial disease care for patients?)

***This interview was designed to help us understand the extent to which the CHABLIS intervention has been implemented, and what has helped or hindered you in doing this. Are there any other important points that you would like to discuss before we close the discussion?***

**Closing and thanks -** check that the participant is still happy for you to use all the information provided and offer the possibility to erase sections of the recording.

Thank them for their time and contribution.

Ensure that they have received a PIS and the CF has been handed back for photocopying.
