## Supplementary material for "Patient and provider perspectives on factors that influence the implementation of a community and hospital care bundle to improve the treatment of patients with peripheral arterial disease in primary and secondary care": Tables 1 and 2 - participant characteristics

| **Patient characteristics** n (%) | Patients  n=11 |
| --- | --- |
| **Age**  41-50 years  51-60 years  61-70 years  >70 years | 1 (9.1)  0 (0.00)  2 (18.2)  8 (72.7) |
| **Gender**  Male  Female | 8 (72.7)  3 (27.3) |
| **Ethnicity**  White British  White Irish  Black/Black British-Caribbean | 9 (81.8)  1 (9.1)  1 (9.1) |
| **Marital status**  Single  Married  Widowed | 2 (18.2)  7 (63.6)  2 (18.2) |
| **Time since diagnosis**  >1 year  1-4 years  5-9 years  10 years + | 3 (27.3)  4 (36.35)  0 (0.00)  4 (36.35) |
| **No. of surgical interventions**  None  Angioplasty  Bypass graft  Amputation | 1  8  4  2 |

Table 1: Patient participant characteristics

| **Provider characteristics** n (%) | Hospital staff  n=8 | GPs  n=6 |
| --- | --- | --- |
| **Age**  21-30 years  31-40 years  41-50 years | 3 (37.5)  5 (62.5)  0 (00.0) | 1 (16.7)  3 (50.0)  2 (33.3) |
| **Gender**  Male  Female | 4 (50.0)  4 (50.0) | 4 (66.7)  2 (33.3) |
| **Ethnicity**  White British  White Other  Asian/Asian British-Indian  Mixed. White & Asian  Chinese  Other | 2(25.0)  1 (12.5)  2 (25.0)  1 (12.5)  1 (12.5)  1 (12.5) | 3 (50.0)  1 (16.7)  2 (33.3)  0 (00.0)  0 (00.0)  0 (00.0) |
| **Core Profession**  Nurse  Vascular surgeon  GP trainee  General practitioner | 1 (12.5)  7 (87.5)  0 (00.0)  0 (00.0) | 0 (00.0)  0 (00.0)  1 (16.7)  5 (83.3) |
| **Grade**  Band 8a  Senior House Officer  Specialty trainee  Speciality registrar  Consultant  GP | 1 (12.5)  2 (25.0)  1 (12.5)  2 (25.0)  2 (25.0)  0 (00.0) | 0 (00.0)  0 (00.0)  1 (16.7)  0 (00.0)  0 (00.0)  5 (83.3) |
| **Time since qualification**  0-2 years  2 years + -5 years  5 years + -10 years  10 years + -15 years | 0 (00.0)  0 (00.0)  3 (37.5)  5 (62.5) | 1 (16.7)  1 (16.7)  2 (33.3)  2 (33.3) |
| **Time in current post**  >6 months  7-12 months  13-24 months  25-36 months  37-48 months  49-60 months  61-75 months  13 years | 5 (62.5)  0 (00.0)  2 (25.0)  0 (00.0)  1 (12.5)  0 (00.0)  0 (00.0)  0 (00.0) | 1 (16.7)  0 (00.0)  2 (33.3)  1 (16.7)  0 (00.0)  0 (00.0)  1 (16.7)  1 (16.7) |

Table 2: Provider participant characteristics
